# Is an integrated model of school eye health delivery more cost effective than a vertical model? An implementation research in Zanzibar

**DOI:** 10.1101/2020.06.30.20143479

**Authors:** Ving Fai Chan, Fatma Omar, Elodie Yard, Eden Mashayo, Damaris Mulewa, Lesley Drake, Mary Wepo, Hasan Minto

## Abstract

**Objective:** To review and compare the cost effectiveness of the integrated model (IM) and vertical model (VM) of school eye health programme in Zanzibar.

**Methods and Analysis:** This 6-month implementation research was conducted in four districts in Zanzibar. Nine and ten schools were recruited into the IM and VM respectively. In the VM, teachers conducted eye health screening and education only while these eye health components were added to the existing School Feeding Programme (IM). The number of children screened and identified was collected monthly. A review of project accounts records was conducted with 19 key informants. The actual costs were calculated for each cost categories, and costs per child screened and cost per child identified were compared between the two models.

**Results:** Screening coverage was 96% and 90% in the IM and VM with 297 children (69.5%) from the IM and 130 children (30.5%) from VM failed eye health screening. The 6-month eye health screening cost for VM and IM was USD 6 728 and USD 7 355. The cost per child screened for IM and VM were USD 1.23 and USD 1.31, and the cost per child identified were USD 51.75 and USD 24.76 respectively.

**Conclusions:** Both models achieved high coverage of eye health screening with the IM being a more cost-effective school eye health delivery screening compared to VM with great opportunities for cost savings.

**Key messages:**

- There is a dearth of information on the actual cost of school eye health (SEH) delivery. This was the first implementation research to review and compare cost effectiveness of the integrated model (IM) versus the vertical model (VM) SEH delivery in Africa.
- VM used 1.2 times more resources per child screened compared to the IM and the cost per child identified in the VM is twice that of the IM, thus the IM is highly cost effective.
- Stakeholders in low- and middle-income settings will be better able to plan for a cost-effective SEH delivery model that suits their contexts and needs using these findings.

## INTRODUCTION

Even though refractive errors can be easily corrected with a pair of inexpensive glasses, worldwide, there are still 12 million children living with vision impairment due to uncorrected refractive errors (URE).[1] Left untreated, refractive error can negatively impact a child’s quality of life,[2] education and future employment[3,4] and can cause significant distress.[5] The common barriers to the uptake of eye services in low and middle-income countries are (i) lack of knowledge and access to available services, (ii) lack of trust and understanding of the treatment outcomes, (iii) cultural and social factors and (iv) parents unaware of the problem if their child does not complain.[6] To address refractive error challenges, many countries conduct school eye health screening programmes because they are simple to conduct, not resource intensive and benefit children with refractive errors. [7]

While there is no formal estimate of refractive errors in Zanzibari children, a case study from the School Health Integrated Programme (SHIP) reported that about 42% of the children in rural Zanzibar communities who needed a pair of glasses did not have them.[8] It was also reported that 90% of vision impairment among Tanzanian children was due to refractive error.[9] Recognising the need to improve public health practices and access to disability-related services,[10] the Ministry of Health in Zanzibar endorsed free spectacle provision for children. Historically, school eye health screening programmes in Zanzibar were implemented in a vertical manner. These were often led by non-governmental organizations and ended abruptly with the cessation of funding as they were not part of the National Health Plan. Hence, in collaboration with a non-governmental eyecare organization, the Ministry of Health of Zanzibar proposed integrating the school eye health programme with its existing school feeding programme (SFP). This provided a valuable opportunity to conduct implementation research to compare the cost-effectiveness of an integrated versus a vertical school eye health programme. We hypothesized that the integrated approach would maximize the limited health resources by utilizing teachers’ time to target both eye health and nutrition within one programme.

### Rationale

Despite mathematical modelling suggesting that screening and correcting refractive error in school children is cost effective in Africa,[11] there is very limited information on the actual costs of school eye health programmes at country level. This makes it extremely difficult to persuade the Zanzibari government to formally integrate school eye health within the National Health Plan, as it needs careful planning and resourcing.[12] Hence, our study aimed to compare (i) the total costs, (ii) the cost per child screened and (iii) the costs per child detected of the vertical model (VM) with the integrated model (IM). We also reviewed the different cost categories to aid realistic budgeting and identify opportunities for cost saving and cost sharing.

## MATERIALS AND METHODS

### School selection

This research protocol was approved by the Zanzibar Medical Research and Ethics Committee (ZAMREC/0001/January/17). The research was implemented in nineteen rural schools from 1 April 2017 to 1 October 2017 in four districts in Unguja (North A and South Districts) and Pemba Island (Micheweni and Mkoani Districts).

All nine schools (approximately 6000 children aged 6 to 13 years old) with an existing SFP were included in the IM. We purposively selected ten schools with no SFP into the IM because the remaining schools in the study area had fewer students. To ensure similar characteristics in the schools enrolled in the two models, we only included rural primary schools which had student school-going rates of approximately 75%, with a similar number of boys and girls and within 5km of the nearest eye centres.

### Description of intervention

#### Phase 1: Training

In April 2017, six master trainers (n=4 for IM and n=2 for VM) were trained in vision screening, recording and referral pathways, and delivering health education to children. As a result of the recommendations made by the local stakeholders, we trained 4 masters trainers for the IM to avoid over-burdening the master trainers and compromised the quality of teachers’ training. More master’s trainers were trained for the project. They subsequently trained 60 teachers (n=30 in both IM and VM) who were appointed by the school head teachers. The training for the IM and VM were a 2-day session and a 1-day session, respectively.

#### Phase 2: Screening and referral

Following their training, teachers returned to their schools and conducted eye health screening from April to September 2017. In the VM, the schools received only the eye health intervention. Teachers conducted the screening, recorded all students screened and identified the students who required follow up. A list of children who were identified with an eye condition was compiled from the student eye screening register. A week after the completion of the eye health screening, children received eye health education using the materials that were developed in collaboration with the Ministry of Health and approved by the Health Promotion Unit. The eye health material package provided to teachers included (i) a Handbook and training manuals on health promotion, (ii) posters for display around the school compound and (iii) an eye health education booklet for use during health education sessions.

For schools in the IM, eye health intervention was added to the SFP. As part of the screening programme, teachers also measured the children’s height and weight (anthropometry measurement). On top of the eye health component, the health education materials included additional information on nutrition, face and hand washing and deworming for children.

#### Phase 3: Monitoring and evaluation

The research coordinator visited the schools monthly to collect the list of children who were screened (eye screening in VM and eye screening and anthropometry in IM) and those who failed screening. Children who failed the eye health screening were referred to a designated vision centre for vision management and children with weight problems were referred to the nearest health centre. The optometrists at the vision centres examined the referred children and managed their eye conditions. The free treatment provided at the vision centre included spectacles and basic eye medication (as per national health policy). The detailed vision screening protocol, and referral criteria are included as supplementary appendices.

The optometrists compiled lists of the children examined and their diagnoses. Cases that could not be managed at the vision centre were referred to Muhimbili Hospital in Dar-es-Salaam for further management. The hospital management was informed of the study so that they were prepared for the increased referrals the screening could create. The description of the IM and VM school eye screening programme is shown in Table 1.

**Table 1:** Description of the school eye screening programmes: integrated and vertical model

|  | **Integrated model** | **Vertical model** |
| --- | --- | --- |
| Number of participating schools | 10 | 9 |
| Number of master trainers trained | 4 | 2 |
| Number of teachers trained | 30 | 30 |
| Number of children enrolled in the schools | 6242 | 5713 |
| Intervention | Eye health, school feeding, deworming and sanitation awareness through teachers | Eye health awareness through teachers training, and Information, Education |
|  | training, and Information, Education and Communication materials | and Communication materials |
|  | Eye health screening by teachers and referral of pupils identified with eye health problems to the nearest vision centre | Anthropometry measurement, eye health screening by teachers and referral of pupils identified with eye health problems to the nearest vision centre<br>Children with weight problem were referred to the nearest health centre |

#### Phase 4: Costing analysis

Project accounts records were reviewed to collect information on resources utilized by the interventions (resource types, numbers and unit costs). Primary data collection was conducted by interviewing key informants from schools, vision centres, districts ministry offices and project sites. Informed consent was obtained from key informants prior to their participation in the study. The key informants were representatives of the Ministry of Health (n=1), Ministry of Education (n=1), District Education Officers (n=4), head teachers (n=4), trained teachers (n=4), optometrists (n=2), project coordinator (n=1), project administrator (n=1), and principal investigator (n=1). The cost categories with their cost components are shown in Table 2.

**Table 2:** Cost categories and cost components for the project

| Cost category | Cost components |
| --- | --- |
| Meetings | Participants' per diem, venue rental, refreshment for one project kick-off meeting, one multi-stakeholder planning meeting and two local project team meetings |
| Staff salaries | Optometrist hourly rate X time spent on examining and managing referred children*, full time staff hourly rate X time spent on eye health activities<sup>§</sup>, teachers' hourly rate X time spent on eye health screening<sup>¥</sup> |
| Outreach activities | Staff per diem, transport, spectacles, eye medication |
| Equipment | Autorefractors, ophthalmoscope/retinoscope sets, slit lamps, trial frames, trial lens |
| Screening kit | Occluder, VA chart, torch, tape measure, vision cards, a height and weight scale, educational books, record books, and posters |
| Monitoring and evaluation (M&E) | Teachers' hourly rate X time spent on monitoring and evaluation activities<sup>α</sup>, per diem, transport<sup>μ</sup>, accommodation, communication<sup>□</sup> |
| Consumables | Spectacles, eye medications |
| Printing | Manuals, branding stickers, posters, and referral forms. |
| Training of trainers | Participants' per diem, venue rental, refreshment |
| Teachers training | Participants' per diem, venue rental, refreshment |
| Baseline and end-line survey | Training costs (venue, per diem, refreshment), data collectors' wages, transport<sup>μ</sup>, accommodation, communication<sup>□</sup> |

*\* Optometrists' time spent on examining and managing the children varied between 7*
*minutes to 60 minutes depending on the complexity of the case at hand*
*§ Full-time staff spent between 25 minutes to 5 hours per school on the eye health activities*
*depending on how well the activities were being implemented*
*¥ Teachers spent 4 to 5 minutes screening one child. Class time used for teaching eye health*
*to students ranged from 10 to 45 minutes per class session.*
*α Teachers' time spent 4 to 12 hours per quarter conducting monitoring and evaluation*
*activities (M&E)*
*μ Means of transportation for M&E included public transport and motorcycle, with the cost*

The collected information was entered, cleaned and analyzed in Excel. Staff time was estimated using the recall method. Costing was done for the period of 6 months (April to September 2017), the actual time taken to implement the two models. Project start-up expenses are considered to assess the project implementation costs but are not included in the costs per child screened and costs per child identified. This rests on the hypothesis that the project start-up costs are only incurred in the first year of the project and absorbed by the relevant ministry costs hereafter. Cost effectiveness of the models were compared in terms of cost per child screened and cost per child identified.

## RESULTS

### Children screened and identified for eye diseases

At the end of the intervention period, 11 987 children were enrolled in the schools, with 6257 children (3127 boys and 3130 girls) in the IM and 5721 children (2960 boys and 2761 girls) in VM. A total of 11 134 (93%) children were screened from April to November 2017, with 5992 children (96%) from the IM and 5142 children (90%) from VM. Of those who were screened, 427 (3.8%) children failed eye health screening, of which 297 (69.5%) were from the IM and 130 (30.5%) from the VM.

### Breakdown of eye health costs in IM and VM

Eye health costs accounted for 60.4% of the total direct cost for the IM. While costs incurred for the training of trainers, teachers training and monitoring and evaluation were similar in the two components, costs incurred for teacher’s salaries, printing and screening kits were higher in the eye health component. (Table 3)

**Table 3:** Total integrated programme costs breakdown

| Cost category | Eye health cost, USD (percentage of integrated programme cost) | Nutrition, deworming and sanitation cost, USD (percentage of programme cost) | Integrated programme cost, USD |
| --- | --- | --- | --- |
| Training of Trainers | 916 (50%) | 917 (50%) | 1 833 |
| Teachers training | 1 650 (49.4%) | 1 690 (51.6%) | 3 340 |
| Monitoring and evaluation | 1 300 (50%) | 1 299 (50%) | 2 599 |
| Teachers' salaries | 545 (69.7%) | 237 (30.3%) | 782 |
| Printing | 504 (66.8%) | 251 (33.2%) | 755 |
| Screening kit | 2 440 (85.3%) | 420 (14.7%) | 2 860 |
| Total Cost (6-month) | 7 355 (60.4%) | 4 835 (39.6%) | 12 170 |

### Cost per child screened and cost per child identified in VM and IM

The eye health cost for the VM and IM was USD 6 728 and USD 7 355, respectively. A total of 5,142 children were screened in the VM and 5,992 children were screened in the IM. Thus, the cost per child screened in the VM was USD 1.31 and IM, USD 1.23. With 130 children identified in the VM and 297 children in the IM, the cost per child identified was USD 51.75 and USD 24.76 respectively. (Table 4)

**Table 4:** Eye health cost per school child in vertical model and integrated model

| Cost category | Eye health cost |  |
| --- | --- | --- |
|  | Vertical model USD | Integrated model USD |
| Training of Trainers | 786 | 916 |
| Teachers training | 1 650 | 1 650 |
| Monitoring and evaluation | 1 114 | 1 300 |
| Teachers' salary | 478 | 545 |
| Printing | 504 | 504 |
| Screening equipment for schools | 2 196 | 2 440 |
| Total Cost (6-month) | 6 728 | 7 355 |
| Total number of children screened | 5 142 | 5 992 |
| Total number of children identified | 130 | 297 |
| Cost per child screened (6 months) | 1.31 | 1.23 |
| Cost per child identified (6 months) | 51.75 | 24.76 |

## DISCUSSION

This study aims to compare the cost effectiveness of delivering school eye health using an IM versus a VM. Our findings show that we were able to screen a high number of children in both models and that the IM is a more cost-effective eye health screening model compared to the VM.

### Children screened and identified for eye diseases

The eye health screening coverages in both models were high (90% in VM and 93% in IM). This screening efficiency was achieved because i) school children are a captive audience and can be reached more easily in schools compared to the general population and ii) there was good coordination in planning and implementing the interventions between the partners of the programme, schools and teachers. We conducted all school screening activities in the first two months of the school semester and before the monsoon season in June because the school attendance rate is highest in this period.

### Costing analysis

Our costing analysis show that the VM used 1.2 times more resources per child screened compared to the IM. The incremental cost of screening the additional 850 children in the IM was USD 627 (USD 0.74 per child). Furthermore, our findings show that the cost per child identified in the IM was about half of the VM, making the IM highly cost effective. A previous study on the cost-effectiveness of school eye screening versus a primary eye health model to provide refractive error services for Indian children by Lester[13] concluded that school eye screening in India is a highly cost-effective method of correcting visual impairment due to refractive errors in school-age children. However, the study used cost per QALY as their cost effectiveness measure and did not describe in detail the difference between the two models and the cost categories of his cost calculation, making it impossible for meaningful comparison between our findings.

We also made a conservative assumption, based on Mariotti and Limburg’s study[14], that the equipment will have a useful life of ten years. However, equipment useful life greatly depends on the existence of a functioning maintenance and repair (M&R) service. In a resource-constrained location such as Zanzibar, where there are no existing M&R services for eye equipment, realistic equipment useful life may be less than ten years. The Ministry of Health has plans to upskill its M&R services to routinely maintain eye equipment, but the timing of this is not yet confirmed.

Training of trainers and teachers training in this pilot project incurred 25.3% of the total project cost. The 1-day (VM) and 2-day (IM) training sessions were needed as these were new approaches for the local implementing personnel. However, investment in intensive training will decrease in subsequent years, and be replaced with less resource-intensive refresher sessions.

The cost-saving opportunities of the IM lie mainly in teacher’s time, training and monitoring activities. The IM has a greater opportunity of reducing resource use as many inputs can be shared with other school-level programmes such as nutrition and other health education programmes.

This pilot project required close and frequent monitoring of the teachers’ eye health screening accuracy. Retraining on site was conducted when required. Hence, we foresee both models incurring less cost as the programme continues and the teachers become more experienced. In the short term (5 years), we project that staff numbers will remain constant with salaries increasing by approximately 4% annually.

One of the long-term aims of the school health programme is to integrate health into the school curriculum, including secondary schools, hence increasing the age intervals of the children screened. Once the integration is achieved, economies of scale will further reduce programme costs. Our findings align with Baltussen et al.’s[14] analysis which showed that while screening a broader-age interval is costlier than a single age-interval, cost per capita can be less because of economies of scale.

All children who required refractive correction were provided with custom-made spectacles regardless of the magnitude of the refractive error, costing between USD 6.8 and USD 14.6 per pair depending on their prescriptions. However, the ability to correct refractive error for many children with ready-made spectacles which are significantly cheaper than custom made spectacles, could reduce programme costs and offer on-the-spot refractive corrections.^[15–17]^

### Limitation

The optometrists’, teachers’ and staff time spent on the activities was determined using the recall method and may have presented a certain level of recall bias. This method might have under- or over-estimated the time spent on the activities. However, we masked the key informants from the model the schools were enrolled into, to minimise response bias.

Baltussen et al.’s[14] findings showed that screening of 11 – 15 years old is the most cost effective in all regions of the world. However, we implemented our operational research among primary school children (6 - 12 years old) because the existing SFPs were in primary schools (IM). To make meaningful cost comparisons between the IM and VM, we implemented the VM in primary schools where the socio-economic profiles were similar to the IM schools.

## CONCLUSION

To the authors’ knowledge, this was the first implementation research that reviewed and compared cost effectiveness of the IM versus the VM school eye health delivery in Africa. While the total implementation cost of the IM was higher than the VM, more children were screened and identified in IM, making it the more cost-effective school eye health delivery model on a per capita basis. Compared to the VM, the IM offers greater cost saving opportunities to achieve long term sustainability. Using these findings, stakeholders in low- and middle-income settings will be better able to plan a cost-effective school eye health delivery model that suits their contexts and needs.

## Data Availability

The data referred in the manuscripts can be provided upon request

## Acknowledgement

We would like to thank Mr Kalam Abushir for his commitment in coordinating the project and Ms Happiness Saronga for data collection.

## Competing interests

None

## Funding

The study was funded by U.S. Agency for International Development Child Blindness Program [grant number PGRD-15-0003-008].

## Notes

### Competing Interest Statement

The authors have declared no competing interest.

### Author Declarations

Zanzibar Medical Research and Ethics Committee (ZAMREC/0001/January/17).

